# Predictive power of SARS-CoV-2 wastewater surveillance for diverse populations across a large geographical range

**DOI:** 10.1101/2021.01.23.21250376

**Authors:** Richard G. Melvin, Nabiha Chaudhry, Onimitein Georgewill, Rebecca Freese, Glenn E. Simmons

## Abstract

The COVID-19 pandemic has exacerbated the disparities in healthcare delivery in the US. Many communities had, and continue to have, limited access to COVID-19 testing, making it difficult to track the spread and impact of COVID-19 in early days of the outbreak. To address this issue we monitored severe acute respiratory syndrome coronavirus 2 (SARS-CoV-2) RNA at the population-level using municipal wastewater influent from 19 cities across the state of Minnesota during the COVID-19 outbreak in Summer 2020. Viral RNA was detected in wastewater continually for 20-weeks for cities ranging in populations from 500 to >1, 000, 000. Using a novel indexing method, we were able to compare the relative levels of SARS-CoV-2 RNA for each city during this sampling period. Our data showed that viral RNA trends appeared to precede clinically confirmed cases across the state by several days. Lag analysis of statewide trends confirmed that wastewater SARS-CoV-2 RNA levels preceded new clinical cases by 15-17 days. At the regional level, new clinical cases lagged behind wastewater viral RNA anywhere from 4-20 days. Our data illustrates the advantages of monitoring at the population-level to detect outbreaks. Additionally, by tracking infections with this unbiased approach, resources can be directed to the most impacted communities before the need outpaces the capacity of local healthcare systems.

## Introduction

Disease outbreak surveillance via wastewater-based epidemiology (WBE) has become an attractive tool for monitoring SARS-CoV-2 at the population and community-levels. WBE has the potential to be especially helpful in localities where access to individual testing is difficult due to limited or sparsely distributed resources. Several studies this year have already demonstrated the realized and potential advantages of WBE for monitoring levels of COVID-19 in communities ^1–4^. In addition to tracking spread, WBE can provide early warning signs of outbreaks and display the efficacy of public health interventions similar to what has been done with polio or herpes viruses ^5^. A study across 7 cities in The Netherlands early during the COVID-19 pandemic showed that fragments of SARS-CoV-2 were first detected in early February when there were no confirmed cases ^6^. Similar studies in Boston, Brisbane, and several other large metropolitan cities around the world have had similar success tracking SARS-CoV-2 in municipal wastewater systems and associating it to the growing number of clinically confirmed cases ^7–9^. These studies highlighted the sensitivity of virus detection in sewage and its potential as a tool to monitor the presence of the virus in large populations. This was especially noteworthy as it is estimated that approximately 80% of COVID-19 infections are mild or asymptomatic ^10^. Given the large number of presymptomatic and paucisymptomatic carriers of COVID-19, wastewater surveillance could be used as an early warning tool to alert to the presence of the virus even if these carriers never receive individual testing. Additionally, clinical data suggested that infected patients can be negative for SARS-CoV-2 in nasopharyngeal samples later in the course of the disease while remaining positive in the gastrointestinal tract, which means a positive signal can be detected much longer from waste ^11^.

With the potential of WBE to monitor infection level trends in regions and communities were testing is limited, the applicability of WBE to these areas has not thoroughly demonstrated. Three major limitations of current WBE SARS-CoV-2 monitoring studies that need to be addressed are a focus on major population centers (≥100,000 persons), lack of longitudinal breadth, and inconsistent use of reporting metrics and normalization across studies ^12,13^. The first limitation, municipality size, does not allow clear understanding of how wastewater surveillance performs when focused on small or large, low population density areas (i.e. rural). There is significant interest in monitoring wastewater in rural communities and congregate living settings (nursing homes, correctional facilities, etc.) as these populations can be particularly vulnerable to low access to healthcare resources ^14,15^. To determine how well wastewater surveillance performs across a range of population levels, this study monitored large (> 100k population), medium (<100,000 and >10,000), and small (<10,000) communities. The second limitation, short sampling periods, prevents making a clear determination of how well wastewater surveillance functions in the long-run as a monitoring/ predictive tool. We sought to address these limitations directly by conducting a longitudinal study of SARS-CoV-2 levels over 20 weeks (May to August 2020) in the municipal wastewater systems of 19 cities in the state of Minnesota, USA. Using 570 sampling points (30 per site) for each of the 19 sites, we formulated a normalized and standardized index for reporting wastewater SARS-CoV-2 levels that takes into account differences in population size and negates the contributions of variation in water flow and treatment facility size. This is the first study to use monitor wastewater SARS-CoV-2 RNA levels in varied populations over a large geographic area (225,000 km^2^). Additionally, we have demonstrated that wastewater SARS-CoV-2 levels can predict changes in clinical cases as far as two weeks into the future. The data presented herein provide evidence that WBE surveillance provides early detection of fast-moving infectious agents like SARS-CoV-2, on both a large and a small scale. However, the power of WBE is largely dependent on normalization of the raw data to reduce the influence of multiple environmental factors that can alter the concentration of pathogens in wastewater.

## Methods

### Wastewater treatment plants

A major goal of this study was to determine the strategy for and effectiveness of large-scale WBE monitoring of disease prevalence across a large region such as a state in the U.S.A. To this end we studied wastewater treatment plants (WWTPs) in the state of Minnesota, recruiting from the state’s major metropolitan area and six surrounding geographical areas designated as North West, North East, Central, South West, South Central, and South East (Figure 1A). To recruit WWTPs, we solicited participation from the 57 WWTP members of the Minnesota Environmental Science and Economic Review Board (MESERB, meserb.org). An enrollment announcement was posted on MESERB’s news web page on April 17, 2020 (http://meserb.org/umd-seeks-facilities-to-participate-in-covid-19-wastewater-study). WWTPs were asked to submit a letter of intent to participate and commit to providing samples through August 31, 2020. Sizes of the population served by each participating WWTP plant, its designed average daily flow rate in millions of gallons per day (MGD), and size of its connected sewer system in kilometers were based on Omana *et al*. (2020) ^16^. Minnesota population numbers were based on United States Census Bureau data (www.census.gov).

**Figure 1.**
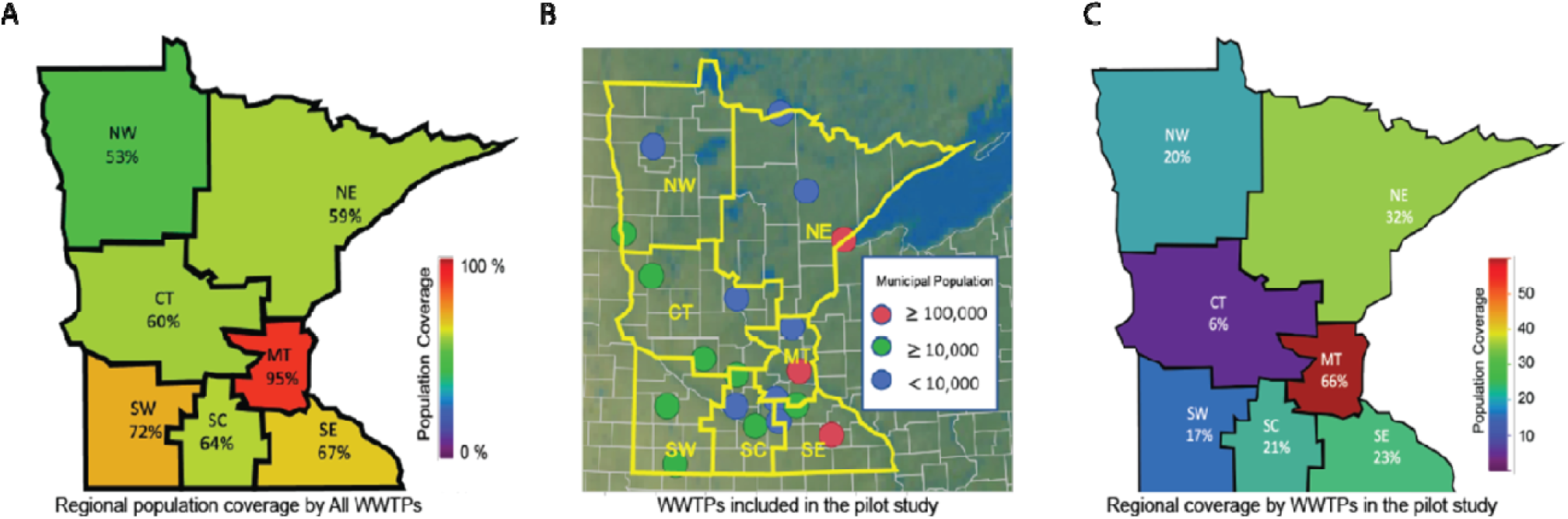
Potential population coverage and study population coverage of wastewater-based epidemiology (WBE) monitoring in the state of Minnesota. A) Existing wastewater treatment plants (WWTPs) can cover 53 to 95% of Minnesota’s regional populations. B) Location and served population sizes of the 19 wastewater treatment facilities that participated in the study. C) WWTPs that participated in the study covered 6 to 66% of regional population.

### Sample Collection and Viral Inactivation

Wastewater (WW) was subsampled from 24-hour composites of total plant influent collected by WWTP personnel. Subsamples consisted of 50 - 500 mL drawn from daily 24-hour total plant intake composites. Collection frequency was once per week for the first month of participation in the study, and then once every two weeks until the end of the study. With the exception of the South East region, where only one WWTP participated, weekly monitoring at the regional level was maintained after the shift in sampling frequency by de-synchronizing submission from the WWTPs. The reduced sampling frequency was necessary for the distribution of limited laboratory staff and resources across the study period during COVID-19 stay at home restrictions in Minnesota.

WW samples were shipped to the laboratory overnight on wet ice. A record of the WWTP’s reported the daily wastewater influent flow rate on the day of collection was included with each shipment. Upon receipt, sample tubes were surface sterilized with 70% ethanol and bleach and then placed into a 60□C water bath for 2 h to Pasteurize the samples. Following Pasteurization, samples were stored at 4□ C until RNA extraction. Storage times were typically 2 h or less.

### Viral Precipitation and RNA Extraction

Solid material was removed from the pasteurized WW samples by filtration through a 0.02µm filter (MilliporeSigma SCGP00525). To precipitate viral particles, 35 mL of filtered WW was combined with 5 mL of 80% PEG-8000, 0.3M NaCl solution in a 50-mL OakRidge centrifuge tube, vortexed to mix thoroughly, and held on ice until centrifugation. Viral particles were pelleted by centrifugation at 12,000 x g for 1.5 h at 4□ C. To extract viral RNA (vRNA), virus pellets were resuspended in Qiazol Lysis Reagent (Qiagen 79306) and incubated 5 min at room temperature. Aqueous and organic phases were separated by addition of chloroform, vortexing and centrifugation at 12,000 x g for 15 min at 4□ C. The aqueous phase was transferred to Qiagen miRNeasy mini spin columns (Qiagen 217004) for RNA purification using manufacturer protocol specifications for micro-RNA isolation. Purified RNA was stored at - 80□ C.

### Quantitative analysis of wastewater viral load

The quantity of SARS-CoV-2 genomic RNA present in the extracted RNA samples was assayed using the CDC emergency use authorization reverse transcription-quantitative polymerase chain reaction (RT-qPCR) protocol and Research Use Only (RUO) reagents. Two amplification targets within the nucleocapsid gene, N1 and N2, were reverse transcribed and amplified using premixed primer/ hydrolysis probes (Integrated DNA Technologies 10006713) and Go-Taq Probe 1-Step RT-qPCR system 2X master mixes with dUTP (Promega A6102). Each 20-µl RT-qPCR contained 5µl of extracted RNA, 1.5µl primer/ probe mix, 10µl Go-Taq master mix, and 0.4µl Go-Script RT mix. Reactions were performed using a Qiagen RotorGene Q thermal cycler with the following thermal profile: reverse transcription at 45□ for 15 minutes; denature and polymerase activation at 95□ for 2 minutes; followed by a 50-cycle two-step amplification profile of 95□ for 5 seconds and 55□ for 30 seconds. Reactions were performed in triplicate (3 measures) within each machine run and positive controls consisting of 200 copies of the amplification target (Integrated DNA Technologies 10006625) and no template controls were included in every run. No amplification was ever observed in the no template controls.

Quantification cycle (cycle threshold, C_T_) was determined using Qiagen Q-Rex software. Quantification of virus copy number was based on standard curves constructed from serial, 10-fold dilutions of positive control plasmid containing the N target gene (Integrated DNA Technologies 10006625). Standard curves for N1 and N2 were estimated using quantile regression on the C_q_-values of the serial dilutions and were then applied to the median of the three measures (N1: Median C_T_ = 40.75 - 3.75 log10 Copy Number; N2: Median C_T_ = 39.78 - 3.08 log10 Copy Number). Log(copies/ Liter of wastewater) was calculated as: Log(copies/ L) = Log[(Copies in reaction)/ 35-ml sample] x 1000 ml/ L].

To standardize qPCR data across the study period and WWTPs, we included reactions that contained primers and hydrolysis probes specific to the Pepper Mild Mottle Virus (PMMoV), a plant virus that is abundant in human feces. PMMoV RT-qPCR reactions contained forward primer 5’-GAGTGGTTTGACCTTAACGTTGA-3’, reverse primer 5’-TTGTCGGTTGCAATGCAAGT-3’ and probe 5’-VIC-CCTACCGAAGCAAATG-BHQ1 and were reverse transcribed and amplified as described above. Standard curves for PMMoV were estimated using quantile regression on the C_q_-values of serial dilutions of Tabasco sauce (McIlhenny Co.) ^17^ and were then applied to the median of the three measures (Median C_T_ = 40.75 - 3.57 log10 Copy Number).

We developed a novel index of WW viral load, Melvin’s Index, that first normalizes the Log(Genome copies/ L) data obtained from the three viral targets and then standardizes based on the normalized PMMoV data (described in Results). The R-code used to calculate Melvin’s Index is available from https://github.com/glennesimmonsjr/dirtywatercooler.

### Recovery of SARS-CoV-2 from wastewater and limit of detection

To determine the efficiency of recovering SARS-CoV-2 from wastewater using our precipitation and RNA purification methods, we spiked known concentrations of heat-inactivated 2019-nCoV-2 (ATCC, VR-1986HK) into sterilized wastewater. Spiked wastewater samples were extracted and assayed using the described protocols. Assays showed recovery efficiencies of greater than 90% when input concentrations were log(6) and log(4) genome copies/ L. Efficiency declined rapidly when input was log(3) genome copies/ L and below. The limit of detection for SARS-CoV-2 virus extracted from wastewater samples was tested using a 10-fold dilution series of heat-killed SARS-CoV-2 in sterilized wastewater. Greater than 95% of qPCR reactions using the CDC N1 and N2 primers/ hydrolysis probes produced a positive detection when SARS-CoV-2 was diluted to as little as 100 copies/ mL (N = 20 for each dilution).

### Correlation of Melvin’s Index with testing data and its predictive value

We tested for correlation between values of Melvin’s Index and confirmed new cases during the study period by calculating the Pearson correlation coefficient between the two variables. To determine the potential predictive window provided by wastewater monitoring, defined as the time between observing an increase in SARS-CoV-2 RNA in wastewater and the appearance of confirmed new COVID-19 positive tests, we successively lagged Melvin’s Index values in 1-day increments and tested for correlation with confirmed new cases. The lag time that maximized correlation between Melvin’s Index and new confirmed cases was taken as the predictive window time in days. Correlation and lag analyses were performed on both statewide averaged Melvin’s Index and on regional Melvin’s Index values. Confirmed new case data was compiled from USAfacts.org ^18^.

### Statistical analysis

All statistical analysis was performed using JMP Pro-15 (SAS Institute, Cary N.C.) or custom R scripts. R scripts are available from https://github.com/glennesimmonsjr/dirtywatercooler.

## Results

### Monitoring SARS-CoV-2 in the wastewater water for the state of Minnesota

As the pandemic spread in the US, our lab became interested in developing WBE monitoring as a way to track viral prevalence at the population-level. Our hope was that a WBE approach could be used to positively impact communities that have been historically underserved by health care systems (low socioeconomic status and/ or rural) ^14,15,19^. Although individual testing for COVID-19 can track virus prevalence in large population centers, major hurdles remained as inequitable access to testing persisted for those who reside in poor and less populated areas ^20^. A major goal of this study was to determine the effectiveness of wastewater surveillance in monitoring the prevalence of SARS-CoV-2 in the large, variously populated, and variously served regions of a U.S. state. Minnesota is the 12th largest U.S. state by area (225,163 km^2^) and 22nd largest by population (5.64 million, the median population of a U.S.A. state is 4.47 million). More than half of the population of Minnesota (58.7 percent) resides in the Minneapolis/ St. Paul metropolitan region and 41.3 percent reside in six geographic regions outside of the metropolitan area. The 856 cities in Minnesota range in population from 429,606 to 8 residents and are served by 586 active, licensed WWTPs. Municipal WWTPs serve 95 percent of Minnesota’s metropolitan population and, depending on region, between 53 to 72 percent of the regional population (Figure 1A). This study was limited to a small number of voluntary participating WWTPs from each region. Fifteen regional and four metropolitan area WWTPs agreed to participate in the study with at least one WWTP participating from each region (Table 1, Figure 1B). Two WWTPs were located in the North West region of Minnesota and represented 20.3 percent of the region’s population, three North East region WWTPs represented 32.5 percent of the region’s population, the three Central region WWTPs represented 6.3 percent of regional population, two South West region WWTPs represented 16.8 percent of the region’s population, four South Central WWTPs represented 21.5 percent of regional population, and one South East region WWTP represented 23.3 percent of that region’s population (Table 1, Figure 1C). The twelve-county metropolitan region of Minnesota was represented by three smaller WWTPs that represented 4.5 percent of the region’s population and one large WWTP that represented 61.4 percent of the population (Table 1).

**Table 1.**
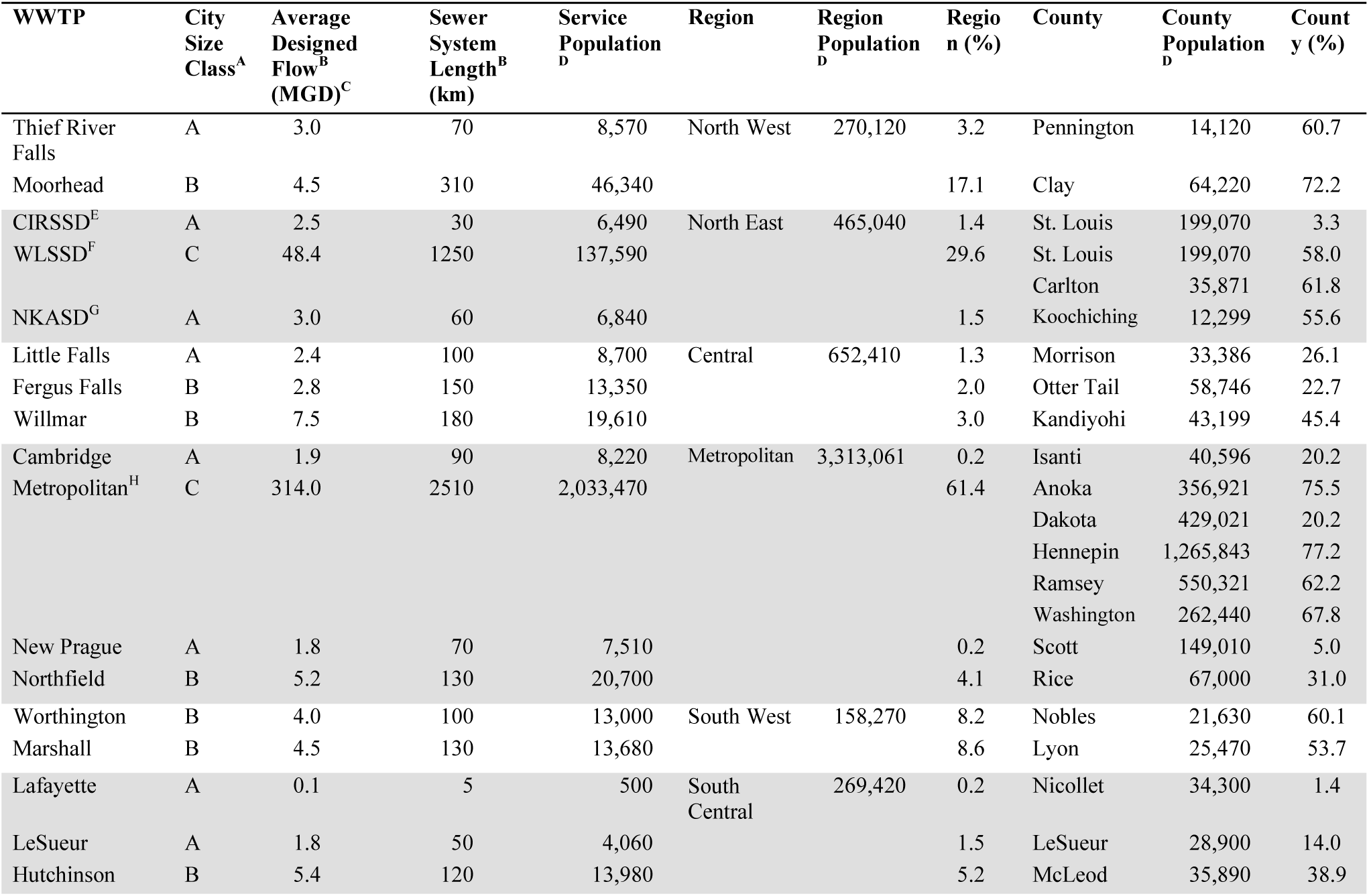
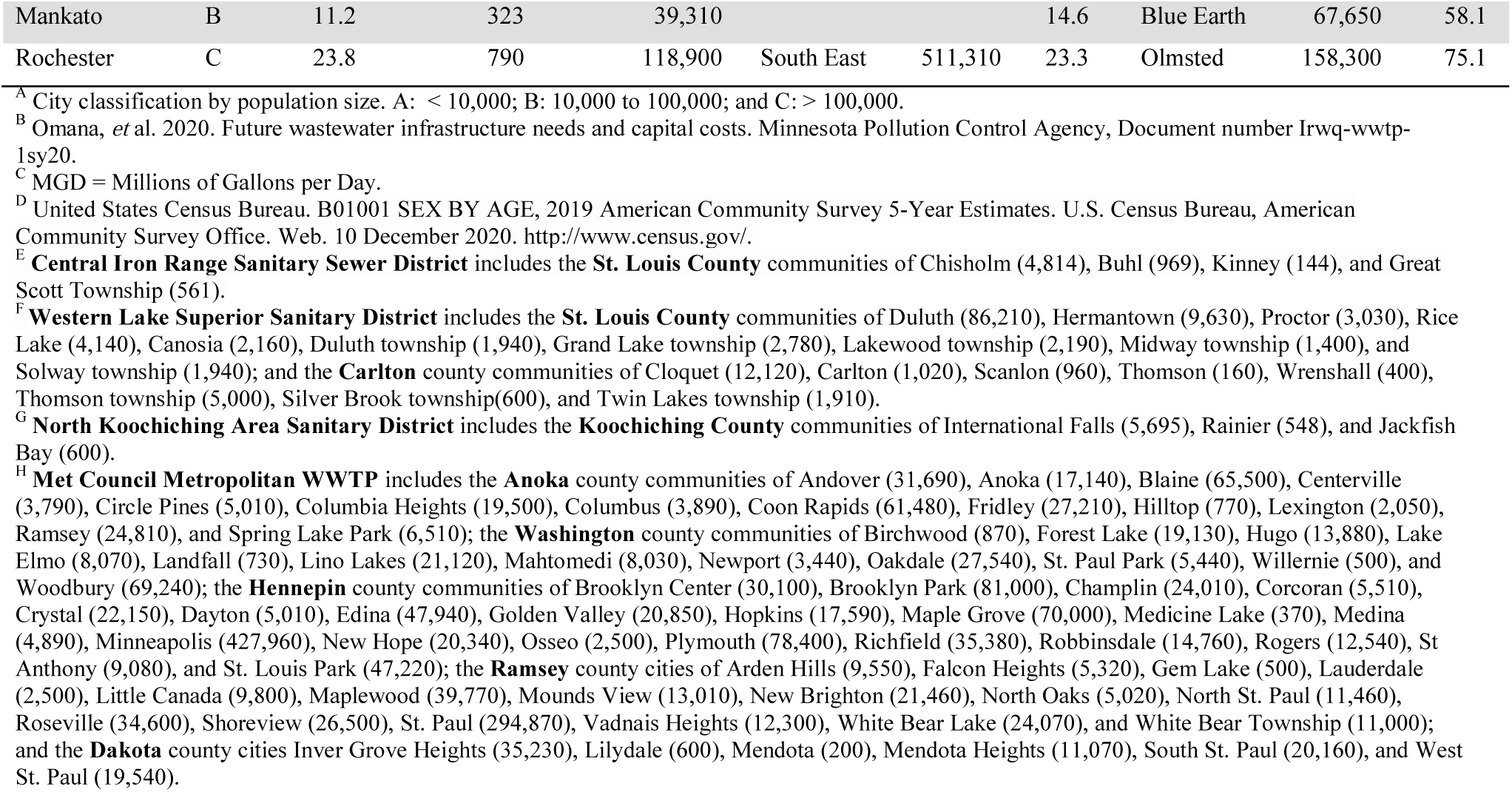
Characteristics of wastewater treatment plants (WWTP) included in the study and their regional population coverage.

The size of the WWTPs was characterized by the size of the population served, the average daily flow rate that the facility was designed to accommodate, and the size of the sewer system. The regional WWTPs served populations ranging from 500 (Lafayette) to 137,590 (WLSSD). Eight WWTPs served a population of less than 10,000 (size class A, median 7,175), eight served between populations 10,000 and 100,000 (size class B, median 16,795), and two served populations larger than 100,000 (size class C, median 128,245) (Table 1, Figure 1B). The largest metropolitan WWTP (Metropolitan) that serves a population of more than 2 million also agreed to participate in the study. Designed average daily flow rate of the regional WWTPs ranged from 0.1 million gallons per day (MGD) to 48.4 MGD with a median of 3.5 MGD. Sewer system length ranged from 5 km to 1,250 km with a median of 110 km. The metropolitan WWTP was designed for an average daily flow rate of 314 MGD and had a sewer system length of 2,510 km. All WWTPs participated for the full study period.

### Effect of environmental factors on detection of virus in wastewater influent

Our primary assumption was that increases in the number of COVID-19 infections in a given city would increase the amount of SARS-CoV-2 RNA detected in that city’s wastewater. However, there are several factors that could affect the concentration of SARS-CoV-2 in the wastewater systems (i.e. flow rate, population, size of WWTP). To better illustrate the magnitude of the differences between each facility, we ranked facilities based on their daily flow rate, Flow rate per capita, and designed flow rate per capita. It is evident that larger cities have larger daily flow rates (Figure 2A). However when facility water flow rates were ranked per capita, the rank order changes dramatically and reveals a concentrating effect on PMMoV level at the largest site (Metropolitan has lowest PMMoV C_T_, Figure 2D), but a dilution effect at the second largest site (WLSSD has fifth highest PMMoV C_T_, Figure 2D). The dilution effect on PMMoV observed for WLSSD was likely due to increased flow from industrial wastewater over the sampling period (Figure 2B). A similar, but less dramatic observation, was seen when ranking facilities based on designed waterflow capacity per capita (Figure 2C). After observing the wastewater influent flow rate characteristics for each facility, we sought to create a normalization and standardization strategy that would account for fluctuations in each system and allow for comparisons between WWTPs. This is an important step toward distinguishing “hot spots” in a large geographic area. To address system variation, we chose Pepper Mild Mottle Virus (PMMoV), an abundant plant RNA virus found in the human diet, as our standard, but we first needed to determine the impact each environmental variable had on PMMoV C_T_-values in wastewater samples.

**Figure 2.**
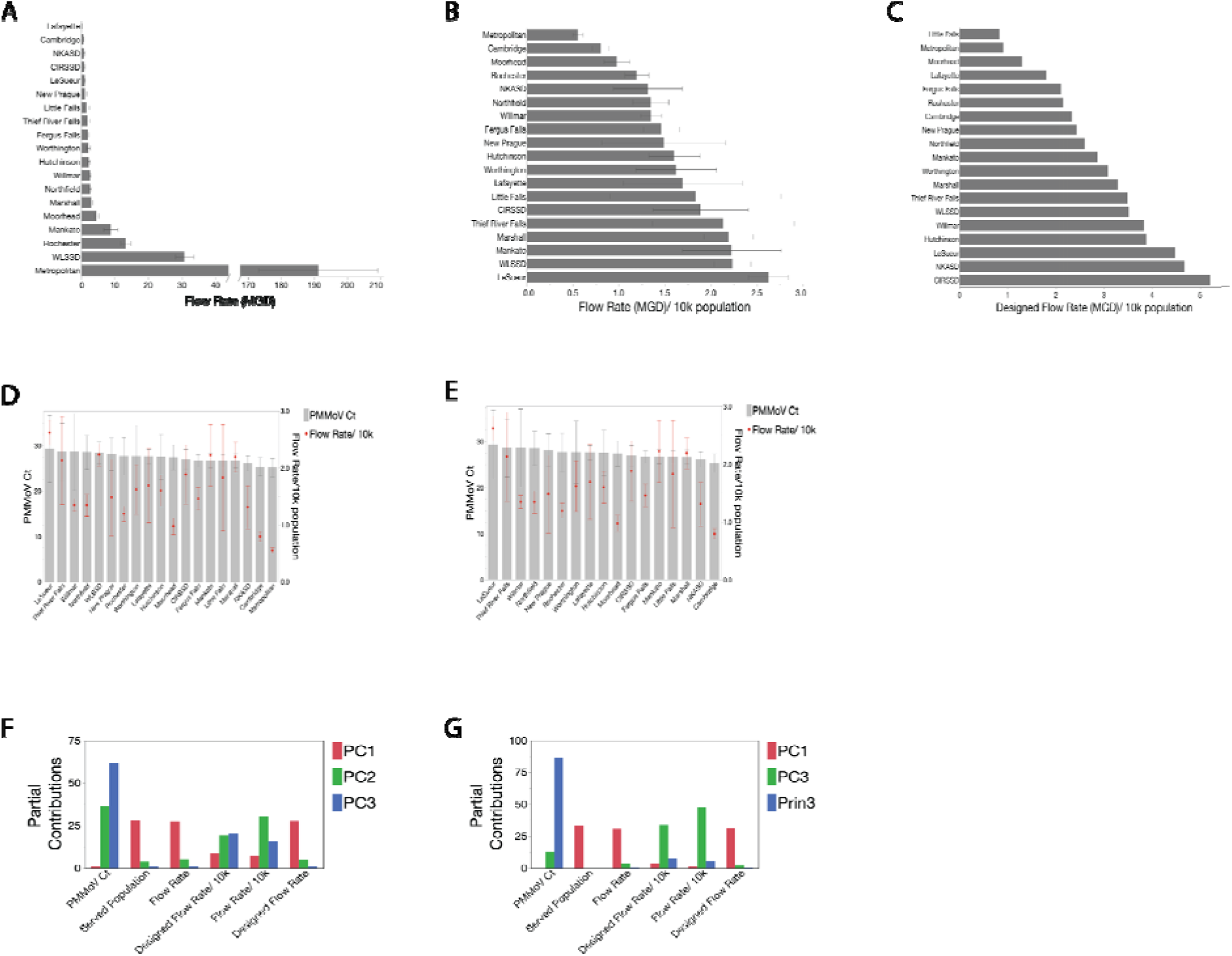
Wastewater treatment facility influent flow rates and served population sizes have little effect on the level of the Pepper Mild Mottle Virus used to standardize SARS-CoV-2 levels. A) Rank order of wastewater treatment facilities based on average daily flow rates. B) Rank ordered wastewater treatment facilities based on daily average flow rates per capita. C) Rank ordered wastewater treatment facilities based on the designed maximum daily flow rate per capita. D) The relationship between ranked order of wastewater treatment facilities based on PMMoV C_T_-values and the respective flow rates per capita. E) The relationship between the ranked order of wastewater treatment facilities based on PMMoV C_T_-values and the respective flow rates per capita, excluding the Metropolitan and WLSSD and facilities. F) Contributions of PMMoV C_T_ and system variables to the three major principal component axes. G) Contributions of PMMoV C_T_ and system variables to the three major principal component axes when the Metropolitan and WLSSD facilities are excluded.

First we tested for a relationship between wastewater influent flow rate variables and PMMoV C_T_-value. PMMoV C_T_-value was relatively stable across the study period and across WWTPs (27.08 ± 3.06, mean ± standard deviation). PMMoV C_T_-value did not differ significantly between WWTPs (one-way ANOVA, F_18, 170_ = 0.69, P = 0.82) despite significant differences between WWTPs in daily flow rate (F_18, 187_ = 1167.05, P < 0.001), designed flow rate (F_18, 187_ = 2.2E+7, P < 0.001), size of served population (F_18, 187_ = 1.0E+14, P < 0.001), flow rate per 10k population (F_18, 187_ = 15.32, P < 0.001) and designed flow per 10k population (F_18, 187_ = 4.0E+15, P < 0.001). The observed stability of PMMoV C_T_ supports its use as a standard and suggests that it could serve as a proxy for population and allow a relative estimation of the infected population size.

It was very important to understand whether variation in system properties were associated with PMMoV levels because the global distribution of PMMoV C_T_ was used to calculate our SARS-CoV-2 level index (Melvin’s index). Use of the global distribution assumes that the level of PMMoV in wastewater is fairly constant within and between populations and that there is a general relationship between PMMoV level and flow rate of the sewer system ^9,21,22^. SARS-CoV-2 level, on the other hand, is expected to vary greatly within and between populations and be subject to the same associations with flow as is PMMoV. If local variables such as flow rate, facility capacity, population size, or the per capita measures of flow and facility capacity strongly affect PMMoV level (and measured PMMoV C_T_) then it may be necessary to use a local rather than a global PMMoV distributions to calculate Melvin’s index.

We observed that WWTPs could be ranked according to their mean PMMoV C_T_-values (Figure 2D). This suggested a potential association with one or more site-specific variables. To better understand which site-specific variables may influence the ranking by PMMoV C_T_, we tested whether flow rate, facility size (designed flow rate), population size, or the per capita measures of flow rate were associated with the PMMoV C_T_-value ranking of WWTPs. When considering all of the participating WWTPs together, per capita flow rate was significantly associated with ranked PMMoV C_T_-values (Kendall’s τ = 0.15, P < 0.01, Table 2, Figure 2D). There was no significant association between PMMoV C_T_ ranking and flow rate, designed flow rate (facility size), size of the served population, or per capita measure of designed flow rate (Table 2).

**Table 2.**
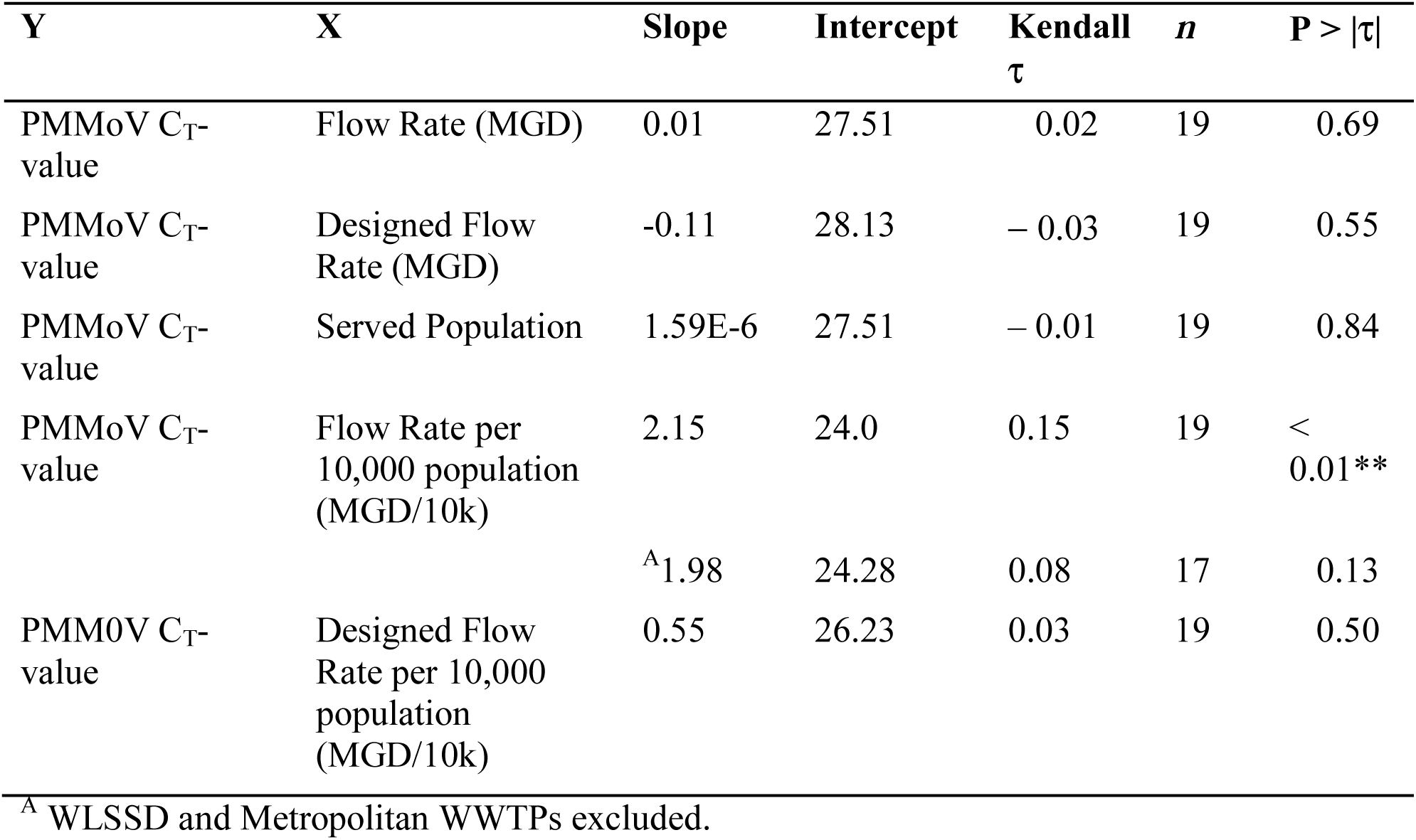
Tests of association between system factors and ranking of PMMoV C_T_-values.

Visual inspection of the per capita flow rate overlaid onto ranked PMMoV C_T_-values (Figure 2D) suggested that their association may be due to the concentration and dilution effects previously noted for two of the larger WWTPs. Metropolitan and WLSSD have strikingly different per capita flow rates (Figure 2B, 2D). The Metropolitan WWTP is a concentrated system that serves a population of 3,454,290 with a per capita flow rate of 0.55 ± 0.5 MGD/ 10k population (mean ± standard deviation) and PMMoV C_T_ of 25.17 ± 2.00. WLSSD is a dilute system that serves a population of 137,590 with a per capita flow rate of 2.23 ± 0.20 MGD/ 10k population (four times higher than that of Metropolitan) and PMMoV C_T_ of 28.44 ± 2.42. Significant association of the per capita flow rate with ranked PMMoV-C_T_ was lost when the WLSSD and Metropolitan WWTPs were removed from the analysis (Kendall’s τ = 0.08, P = 0.13), but not when either was removed individually (τ = 0.12, P < 0.05 and τ = 0.11, P < 0.05, respectively when Metropolitan or WLSSD were excluded) (Figure 2E, Table 2). Although rankable, PMMoV C_T_, did not differ significantly between WWTPs. This finding demonstrated that even large local differences in flow rate have a small effect on PMMoV C_T_ and supported both the use of PMMoV as a standard and the global PMMoV distribution in the calculation of Melvin’s index.

To further test for relationships between PMMoV C_T_-value, flow rate, served population size, and designed flow capacity of the WWTP, we performed a principal component analysis (PCA) on the correlation between variables. PCA combines variables that are associated with each other onto a common axis termed a principal component (PC) and quantifies the contribution of each variable to that axis. The strength of association between variables can be inferred from their respective contributions to the PC axes that they share (Figures 2F and 2G). PCA analysis that included all WWTPs showed that 74.2 percent of the variation between all variables was described by two principal components (PC1 and PC2) (Figure 2F and Table 3). A further 15.3 percent of the variation was explained by PC3. PMMoV C_T_ contributed mainly to PC2 and PC3 (36.8 and 61.6 percent, respectively) with negligible contribution to PC1 (Figure 2F). PCA showed a modest association between PMMoV C_T_, per capita flow rate, and per capita designed flow rate through their common contributions to PC2 (30.4 and 19.2 percent, respectively) and PC3 (15.4 and 20.0 percent, respectively) (Figure 3F). Served population, flow rate, and designed flow rate made negligible contributions to PC2 and PC3 indicating very weak association with PMMoV C_T_.

**Table 3.**
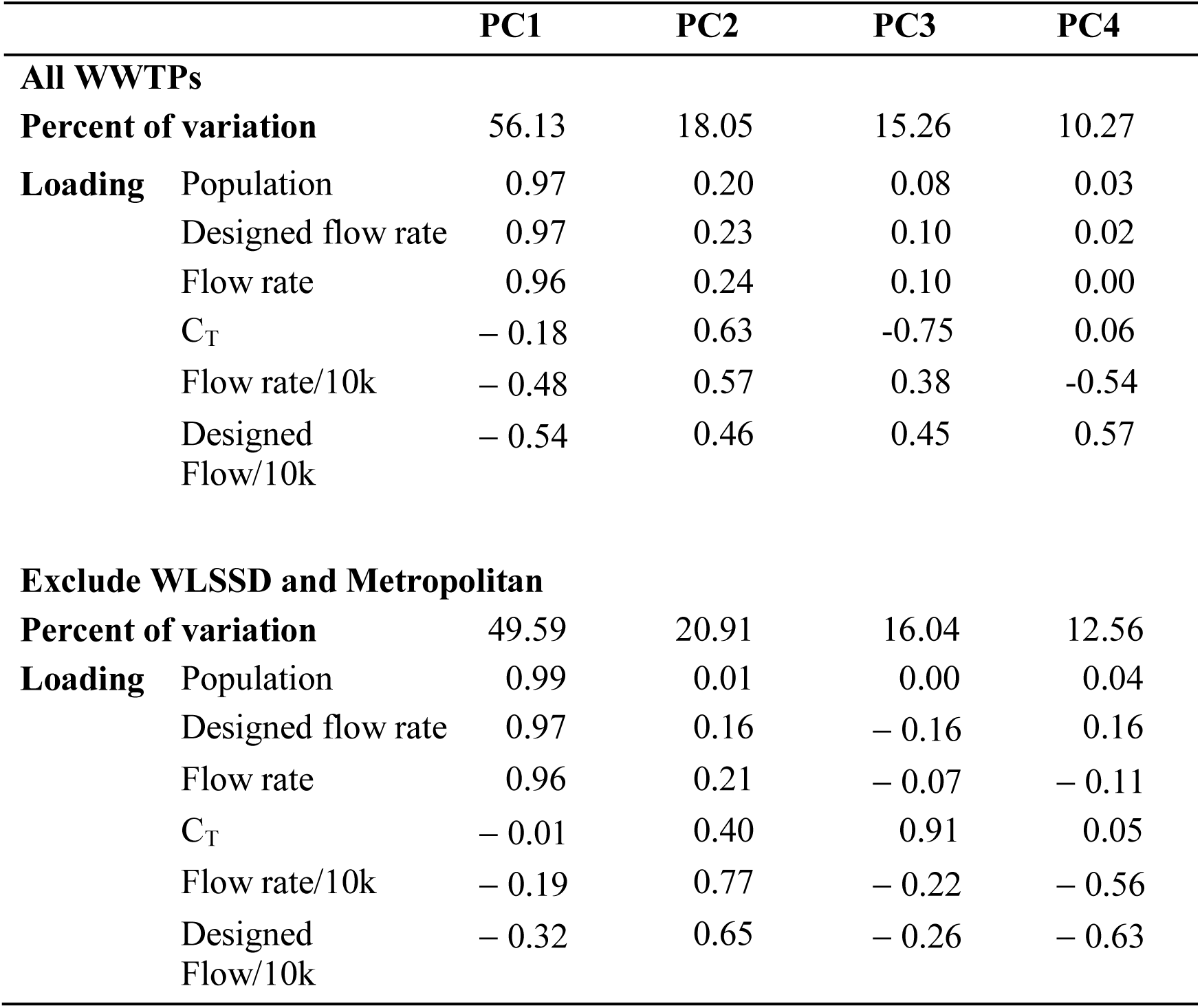
Principal components analysis on correlations. Percent of variation contained by each principal component and the loading for each variable.

**Figure 3.**
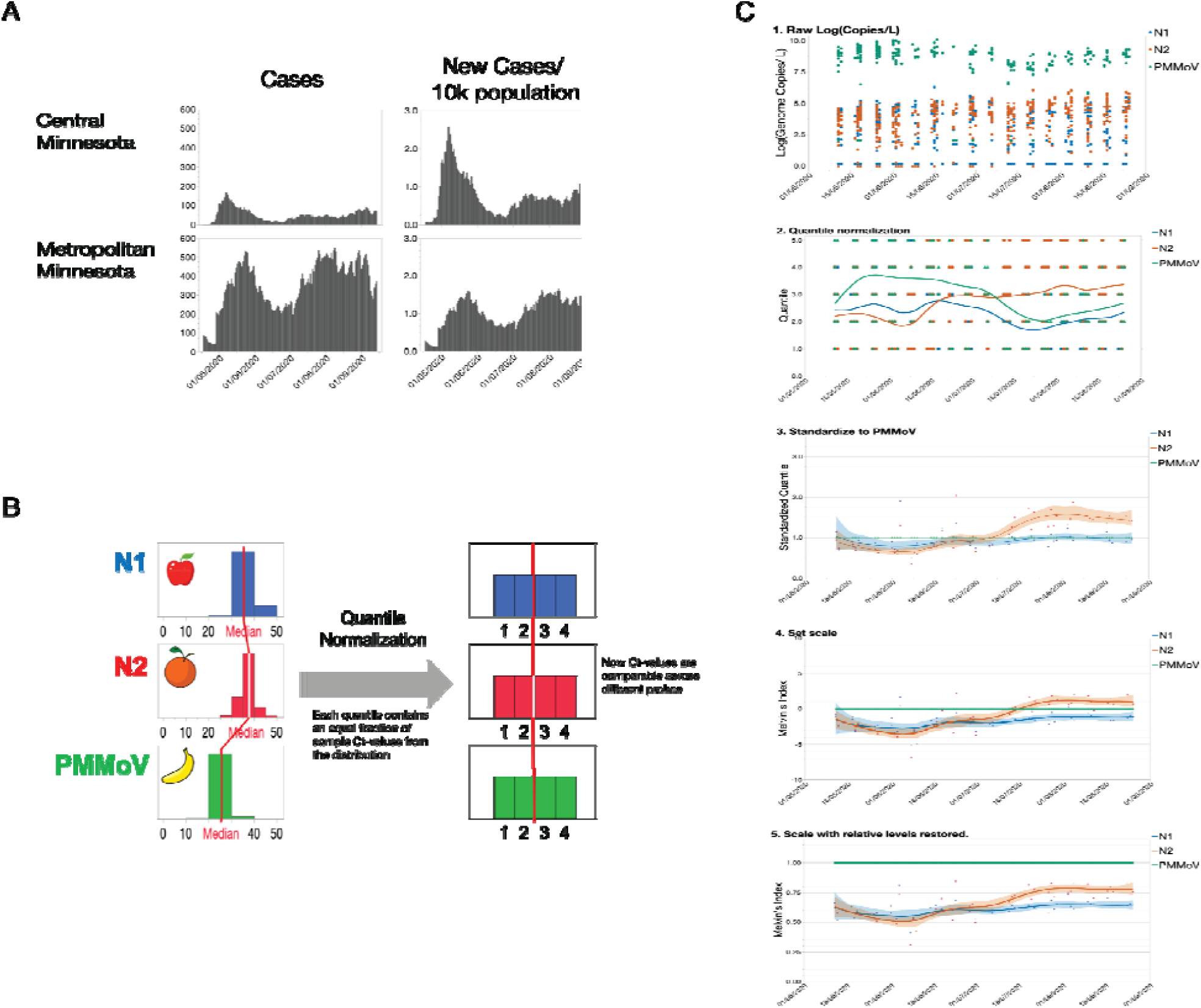
Melvin’s Index of wastewater viral load incorporates quantile normalization, standardization, and intuitive scaling for the tracking of SARS-CoV-2 in wastewater. A) The effect of per capita normalization on interpretation of a comparison of new COVID-19 cases between two regions of Minnesota. Normalization shows that disease prevalence was higher in the more sparsely populated Central region than in the Metropolitan region during early May 2020. B) Graphical depiction of the quantile normalization process. C) Step-by-step conversion of raw Log(Genome copies/ L) data to the normalized and standardized Melvin’s Index.

**Figure 4.**
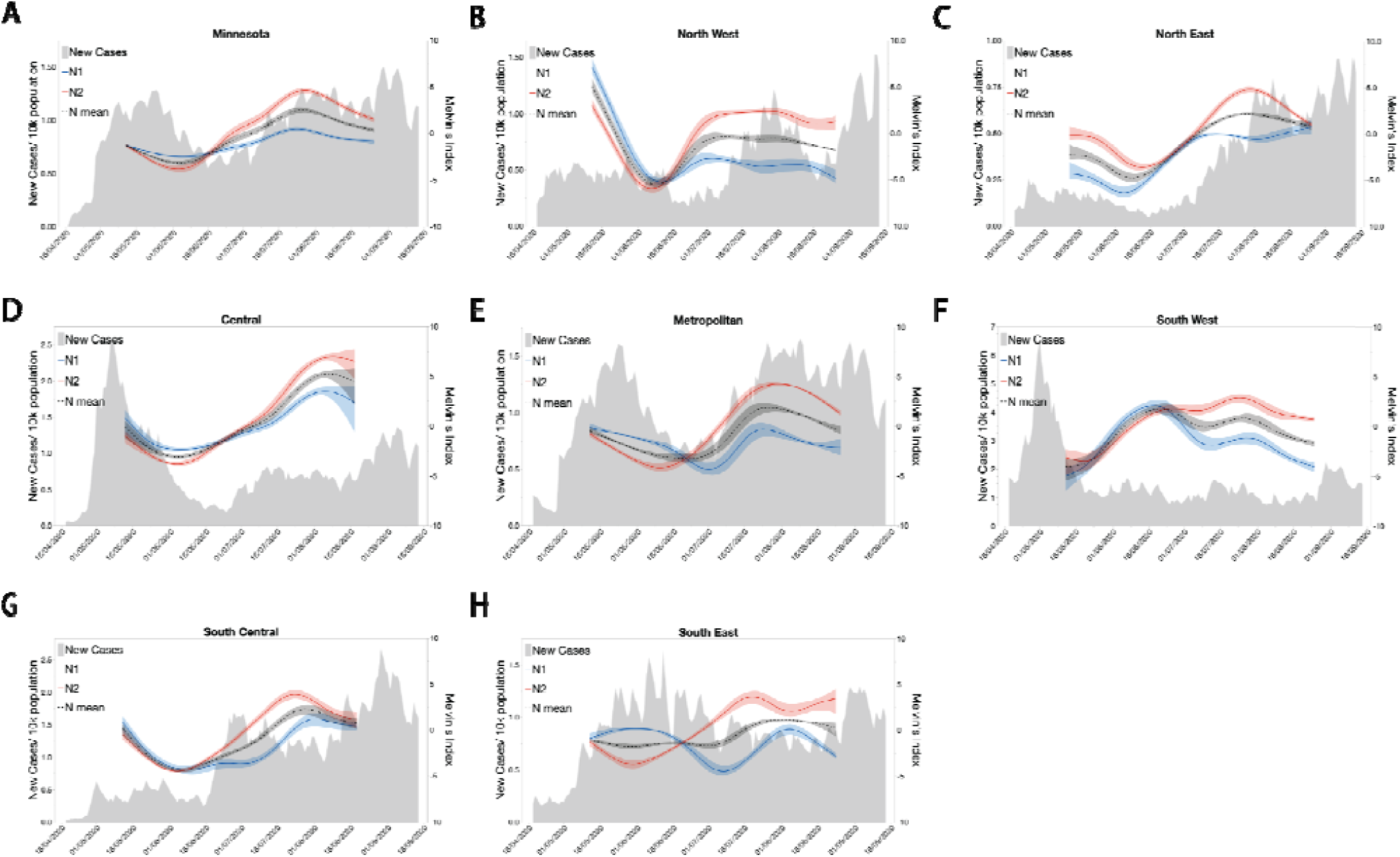
Trends for indexed wastewater SARS-CoV-2 levels across the state show a similar pattern to that of new cases per capita with a temporal offset. A) New clinical cases of COVID-19 overlayed by the indexed SARS-CoV-2 N1, N2, and the mean of N1 and N2 data from RT-qPCR analysis of wastewater samples collected across the state of Minnesota from May to September. B, C, D, E, F, G, H) New clinical COVID-19 cases overlayed by indexed SARS-CoV-2 N1, N2, and the mean of N1 and N2 data from RT-qPCR analysis of wastewater collected from different regions of the state of Minnesota.

**Figure 5.**
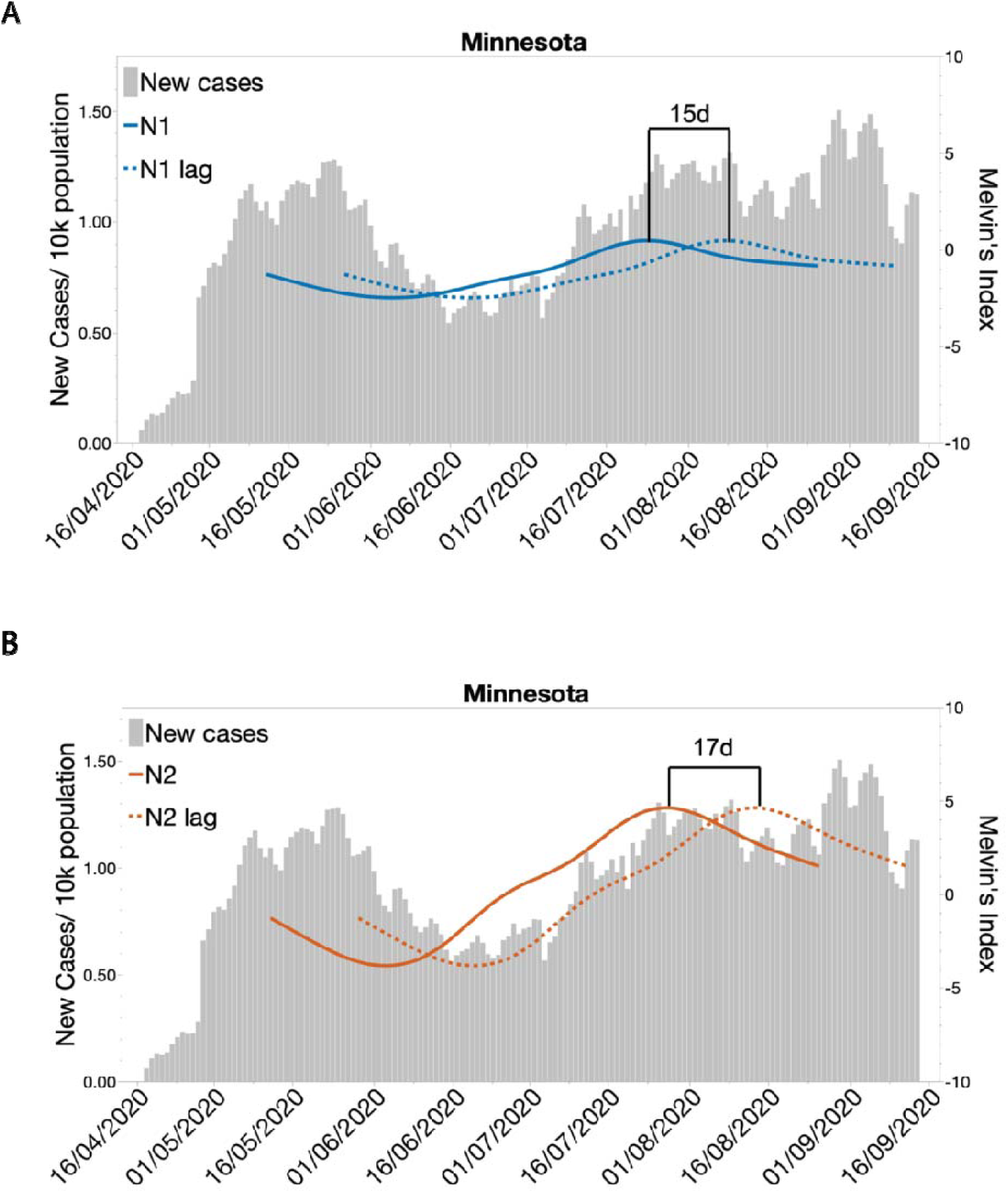
Indexed SARS-CoV-2 levels in municipal wastewater predict new clinical cases in Minnesota. A) Correlation and lag analysis of wastewater SARS-CoV-2 N1 levels with new clinical cases of COVID-19 for the state of Minnesota. B) Correlation and lag analysis of wastewater SARS-CoV-2 N2 levels with new clinical cases of COVID-19 for the state of Minnesota. ***, indicates p < 0.001

To confirm that the modest association between PMMoV C_T_ and per capita flow rate was primarily due to the WLSSD and Metropolitan WWTPs, we removed both facilities from the analysis (Figure 2G and Table 3). In agreement with the earlier tests, the contribution of PMMoV C_T_ to PC2 (the axis that captured most of the per capita flow variation) decreased to 13 percent and its contribution to PC3 (the axis that captured most of the PMMoV C_T_-value variation) increased to 86.6 percent. Contributions of the per capita measures of flow and designed flow to PC2 were increased (47.5 and 33.9 percent, respectively) and their contribution to PC3 was decreased (5.2 and 7.3 percent, respectively). Population size served, flow rate, and designed flow capacity contributed negligibly to PC3. Likewise, PMMoV C_T_ was a small contributor to PC2 (per capita flow) and a negligible contributor to PC1 (flow rate, designed flow rate, and population size). Taken together, these results showed that flow rate and population size have negligible to low-modest association with PMMoV C_T_ and confirmed our previous conclusions that PMMoV was a suitable standard, and that the global PMMoV distribution could be used in our calculation of Melvin’s index.

### Data normalization, standardization, and creation of a wastewater viral load index (Melvin’s Index)

A goal of this study was to provide communities with a readily interpretable picture of changes in viral prevalence through WBE monitoring. Most commonly, wastewater viral load has been reported as qPCR C_T_ or as the logarithm of genome copies per liter of wastewater ^1,2,6,7,9,23–27^. While those measures hold meaning for experts, they can be opaque to the communities and officials who use the information. Data that are reported without appropriate normalization can make it difficult to discern the relative magnitude of the disease prevalence over time or between localities. For example, normalizing the number of new cases per capita showed that in early May 2020, SARS-CoV-2 was more prevalent in Central Minnesota than in the Metropolitan region (Figure 3A). We created a normalized and standardized index of wastewater viral load (Melvin’s Index) to provide a simplified, scaled value for benchmarking and tracking virus levels.

To create the index, we needed to address two issues of qPCR testing. The first issue was that of normalizing between different primer/ probe sets. To address the issue, we applied quantile normalization (Figure 3B and 3C). A second issue was that of samples which did not produce a reliably detectable amplification signal within 50 PCR cycles. Samples that do not amplify are often termed “negative tests” and are excluded. Exclusion of data may create a bias that leads to overestimation of virus load. To preserve the information of negative tests, we assigned a C_T_-value of 51 to samples that did not amplify. Inclusion of the C_T_ = 51 data could create a bias leading to underestimation of viral load. To reduce bias, we excluded the C_T_ = 51 data points from the quantile normalization step. However, following normalization, the C_T_ = 51 data points were added back to the first quantile of the normalized data.

To calculate the index value, the mean of the two or three C_T_ measures was calculated for each WWTP, sample date, and target gene. When the mean of the three C_T_-values equaled 51 for a target gene, the data point was temporarily excluded from quantile normalization for that target gene. The quantity Log (Genome copies/ L) was calculated for the remaining data using target gene specific standard curves. Quantile normalization of the Log(Genome copies/ L) data was performed separately for the N1, N2, and PMMoV target genes. For each target gene, the distribution of Log(Genome copies/ L) was divided into 5 equal bins that each contained 20% of the data, with bin 1 containing the bottom 20 percent of the data, and bin 5 containing the top 20% of the data. The data falling into each bin was reassigned the value (*Q5*) of its respective bin (*Q5* = 1, 2, 3, 4, or 5). All C_T_ = 51 data points were then added back to the data set and assigned to bin 1. Following normalization, the data set consisted of *Q5*-normalized values of the target genes (*N1_Q5, N2_Q5*, and *PMMoV_Q5*) for each WWTP and sample date. N1 and N2 data were standardized to PMMoV by dividing their *Q5*-normalized values by that of PMMoV. The standardized data (*StdQ5*) were natural log transformed to produce linear scaling and multiplied by 10/*Ln(*5) to set the scale range from -10 to 10. We call the values produced by this last transformation “Melvin’s Index” (MI) (Figure 3C).

Because MI-values are centered on the median MI-value for each target gene, the relative levels of virus can be compared over time within N1, N2, and PMMoV, but not between target genes or viruses. We were concerned that the loss of relative level might cause misinterpretation by a non-expert audience. For example, in Figure 3C, the MI-value of N2 rises above that of the highly abundant PMMoV virus. To prevent misinterpretation, we created a modified Melvin’s Index (mMI) that maintains relative level relationships. To calculate the modified Melvin’s Index, the *StdQ5* is multiplied by the ratio of the median Log(Genome copies/ L) for the N gene of interest to that of PMMoV

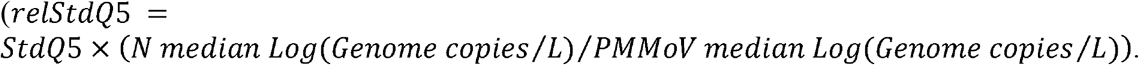

Finally, the *relStdQ5* is natural log-transformed and then scaled to set mMI(PMMoV) equal to 1.0 by adding the natural number (*e*) to Ln(RelStdQ5) and then dividing the sum by *e (Ln(RelStdQ5 + e)/e)* (Figure 3C). R code that uses C_T_-values as input is available from https://github.com/glennesimmonsjr/dirtywatercooler.

### Indexed SARS-CoV-2 levels in wastewater show similar trend to new clinical COVID-19 cases

Our laboratory collected wastewater for over five months from 19 municipal wastewater sites, acquiring approximately 570 sample points. The clinical data for the state showed that SARS-CoV-2 virus was present in late Spring and then began to decrease until around mid-July, but then it steadily increased through August (Figure 2A, grey shading). Similar trends were observed for indexed wastewater SARS-CoV-2 RNA in each region across Minnesota, with the exceptions of the South East and South West regions (Figure 2F and 2H). Interestingly, indexed wastewater SARS-CoV-2 RNA data pooled for the entire state had a similar pattern to clinical cases of COVID-19 around the same period, except it appeared to be slightly offset (Figure 2A). Of particular note, the levels in wastewater samples (indexed SARS-CoV-2) appeared to increase by mid-June, preceding the apparent increase in new clinical cases. Again, this trend was recapitulated in most of the other regions of Minnesota, expect for the South West and South East regions (Figure 2 B-H).

### Correlation of wastewater testing with population testing data and its predictive value

To test the potential predictive value of wastewater we performed correlation and lag tests against Minnesota Department of Health confirmed new case data over the study period. Statewide averaged Melvin’s Index values showed low but significant correlation with statewide new confirmed cases. Pearson correlation coefficient comparing N1 to new cases was 0.38, that comparing N2 to new cases was 0.38, and that comparing the mean of N1 and N2 to new cases was 0.39. For statewide averaged data, lag analysis showed that wastewater predicted confirmed new cases by 15d for N1 (*r*_107_ = 0.73, P < 0.001) and 17d (*r*_107_ =0.77, P < 0.001) for N2. The predictive window when Melvin’s Indices for N1 and N2 were averaged was 17d (*r*_107_ =0.68, P < 0.001). When regions were considered, lag analysis showed less predictive power. Central Minnesota had greatest agreement between N1 and N2 predictive windows (N1: 23d, *r*_98_ = 0.66, P < 0.001 and N2: 23d, *r*_98_ = 0.78, P < 0.001). The North East region had greatest correlation for both N1 and N2 Melvin’s Index values with confirmed new cases (Table 4).

**Table 4.**
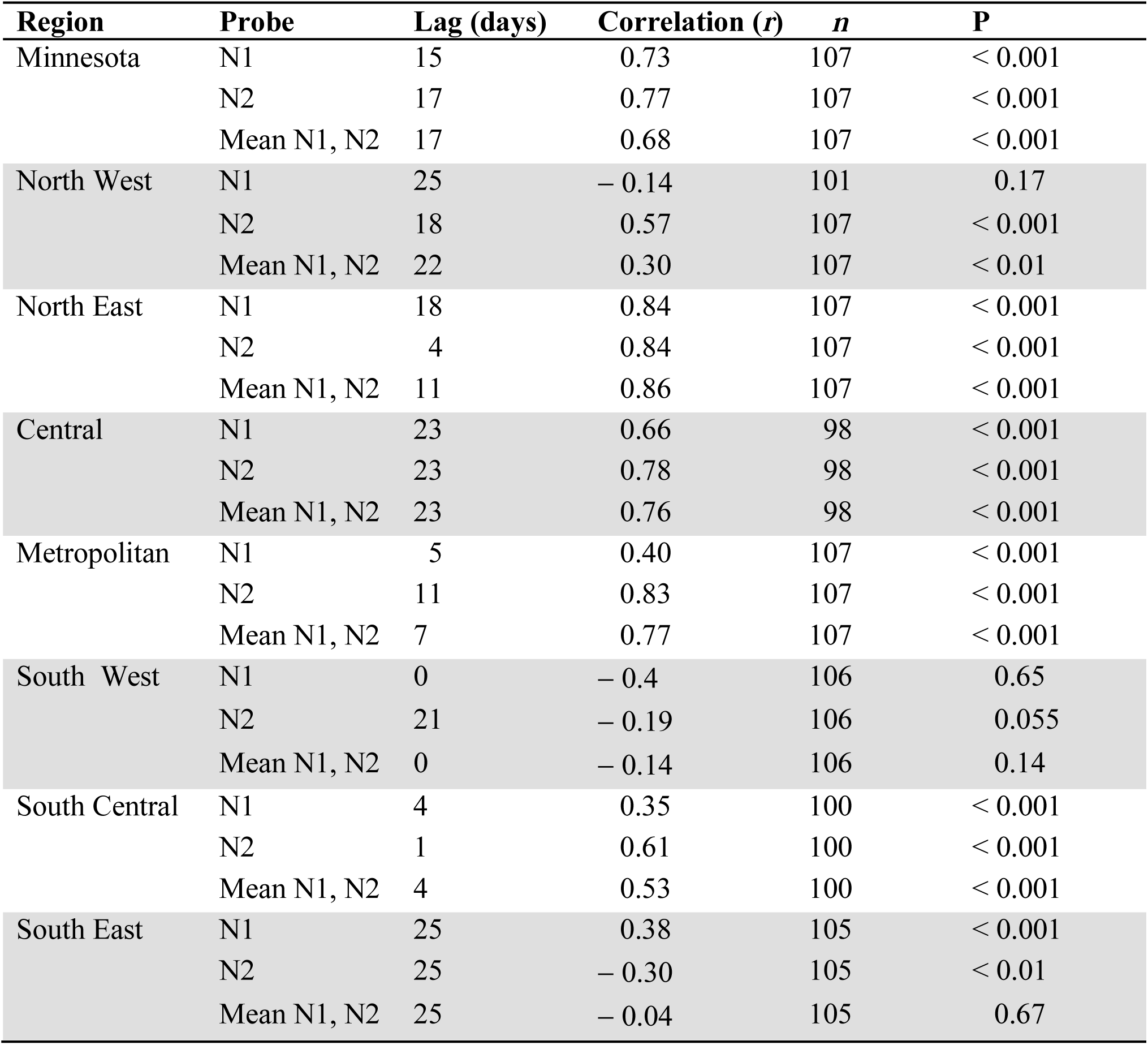
Lag analysis comparing Melvin’s Index to new COVID-19 cases/ 10k population.

| **Region** | **Probe** | **Lag (days)** | **Correlation (*r*)** | ***n*** | **P** |
| --- | --- | --- | --- | --- | --- |
| Minnesota | N1 | 15 | 0.73 | 107 | < 0.001 |
|  | N2 | 17 | 0.77 | 107 | < 0.001 |
|  | Mean N1, N2 | 17 | 0.68 | 107 | < 0.001 |
| North West | N1 | 25 | − 0.14 | 101 | 0.17 |
|  | N2 | 18 | 0.57 | 107 | < 0.001 |
|  | Mean N1, N2 | 22 | 0.30 | 107 | < 0.01 |
| North East | N1 | 18 | 0.84 | 107 | < 0.001 |
|  | N2 | 4 | 0.84 | 107 | < 0.001 |
|  | Mean N1, N2 | 11 | 0.86 | 107 | < 0.001 |
| Central | N1 | 23 | 0.66 | 98 | < 0.001 |
|  | N2 | 23 | 0.78 | 98 | < 0.001 |
|  | Mean N1, N2 | 23 | 0.76 | 98 | < 0.001 |
| Metropolitan | N1 | 5 | 0.40 | 107 | < 0.001 |
|  | N2 | 11 | 0.83 | 107 | < 0.001 |
|  | Mean N1, N2 | 7 | 0.77 | 107 | < 0.001 |
| South West | N1 | 0 | − 0.4 | 106 | 0.65 |
|  | N2 | 21 | − 0.19 | 106 | 0.055 |
|  | Mean N1, N2 | 0 | − 0.14 | 106 | 0.14 |
| South Central | N1 | 4 | 0.35 | 100 | < 0.001 |
|  | N2 | 1 | 0.61 | 100 | < 0.001 |
|  | Mean N1, N2 | 4 | 0.53 | 100 | < 0.001 |
| South East | N1 | 25 | 0.38 | 105 | < 0.001 |
|  | N2 | 25 | − 0.30 | 105 | < 0.01 |
|  | Mean N1, N2 | 25 | − 0.04 | 105 | 0.67 |

## Conclusions

In this study, we have demonstrated the effective application of wastewater surveillance to monitor infectious disease outbreaks across a large and variously populated geographic area. We have improved the interpretation and communication of the results of wastewater monitoring by converting cryptic raw SARS-CoV-2 qPCR data into a pellucid, normalized, and standardized index value (Melvin’s Index) that correlates well with the appearance of new COVID-19 cases. Melvin’s Index accounts for population variation and provides a means of directly comparing viral prevalence between sites that are dispersed across large geographic areas. Further, we have shown that, by using our method of indexing SARS-CoV-2 RNA levels, wastewater monitoring can presage changes in new clinical cases by as much as two weeks (15-17 days) in both rural and in large metropolitan areas.

Interestingly, wastewater SARS-CoV-2 RNA data from two regions of Minnesota (South East and South West) did not strongly correlate with clinical case trends (Table 4). This finding likely reflects circumstances unique to these two regions such as differences in COVID-19 testing, implementation of public health mitigation strategies, or other variables that are not present in other parts of the state. Of note, the South East region of Minnesota is a major manufacturing, agriculture product processing, and healthcare hub for the state, all of which may have impacted the frequency of testing and geographical tagging of testing results. The South West region of Minnesota experienced large outbreaks of COVID-19 immediately prior to the beginning of our study which may have modified population behavior and testing frequency.

Lack of, or weaker correlation between cases and wastewater SARS-CoV-2 RNA in other parts of the state may be an indicator of disparities in health services between metropolitan and regional communities and between communities within a region. For example, regions contain counties in which the population is concentrated in one or two urban areas as well as counties in which the population is dispersed in several rural communities. Several lines of evidence have shown that low-income, underserved, and less populated communities have experienced additional burdens during this pandemic that may be reflected in the low correlation between wastewater and confirmed cases ^14,15,20^. However, in general, health outcomes data for southern Minnesota indicate that it is a relatively healthy part of the state ^18,19,28^. How disparities affect performance of wastewater testing should be explored in future studies. Nevertheless, based on the data that we have generated in this study, we are confident that indexed measures of wastewater SARS-CoV-2 RNA levels accurately depict population-wide changes in disease prevalence. Future studies should focus on determining sampling strategies that maximize population coverage.

The dynamic nature of our sampling, in terms of both geographic and population size, provides additional evidence that wastewater surveillance is an effective and powerful public health implement for tracking disease outbreaks in locations deficient of readily available testing. Additionally, this study serves as a step toward proof-of-concept studies that employ smaller, neighborhood-scaled sampling within larger population centers. In this study we have shown that wastewater monitoring can be effectively deployed in populations as small as 500. Overall, our study demonstrates the real potential of wastewater surveillance in the current pandemic and sets the stage for implementing a large network of surveillance sites for monitoring and prevention of future outbreaks across large geographic areas. Expansion of these demonstrably successful strategies of wastewater monitoring to include high throughput sequencing technology for the surveillance of emerging SARS-CoV-2 strains and other emerging pathogens should be a high priority for future research.

## Data Availability

The data that support the findings of this study are available from the corresponding author, GES, upon reasonable request.

https://github.com/glennesimmonsjr/dirtywatercooler

## Acknowledgements

We wish to thank the Minnesota Environmental Science and Economic Review Board and all the affiliated wastewater treatment facilities and operators for their dedication to this work. The work was supported by the UMN Faculty Driven Award (GES) and the CTSI Pre-K Discovery Scholar Award (GES). The authors thank S. Champeau, D. Wisniewski, K. Andresen, T. Mosley and S. Sundeen who all provided technical support and commented on manuscript drafts.

## Author contributions

RGM, NC, OG, RB, and GES wrote, edited, and proofread the manuscript. RGM, RB performed statistical analysis. RGM and GES designed and carried out this study

